# Protease-activated receptor-1 expression in cytotrophoblasts and platelet-fibrin thrombus formation increase in placenta accreta spectrum

**DOI:** 10.64898/2026.01.15.26344241

**Authors:** Murasaki Aman, Toshihiro Gi, Nobuyuki Oguri, Eriko Nakamura, Kazunari Maekawa, Sayaka Moriguchi-Goto, Yuki Kodama, Shinji Katsuragi, Yujiro Asada, Yuichiro Sato, Atsushi Yamashita

## Abstract

**Background:** Placenta accreta spectrum (PAS) is characterized by abnormal trophoblastic invasion into the uterine myometrium and is a cause of postpartum hemorrhage and maternal death. Protease-activated receptor-1 (PAR-1) promotes various cellular actions, including invasion. Here, we analyzed the expression of PAR-1, platelet antigen, and fibrin in PAS.

**Methods:** We analyzed 49 PAS cases (placenta accreta vera [accreta vera], 31 cases; placenta increta [increta], 8 cases; placenta percreta [percreta], 10 cases, classified by the degree of placental villous invasion) and 33 control cases. We immunohistochemically examined the expression of PAR-1, platelet glycoprotein (GP) IIb/IIIa, and fibrin.

**Results:** The frequency of previous cesarean section was higher in the increta and percreta groups than in the control and accreta vera groups. PAR-1 expression in placental villi was weak and limited in extent in control cases, whereas immunoreactivity and staining density increased in increta and percreta. Immunofluorescence revealed PAR-1 expression in cytotrophoblasts of placental villi and in aggregated platelets. PAR-1 expression scores in cytotrophoblasts increased significantly with the degree of villous invasion (accreta vera, increta, percreta) compared with controls. The immunopositive areas for GPIIb/IIIa and fibrin were significantly larger in PAS groups than in controls. Furthermore, the immunopositive areas for platelets and fibrin were positively correlated with the PAR-1 expression score.

**Conclusion:** These results indicate that PAR-1 may play a role in placental villous invasion and that a thrombogenic placental environment may influence PAR-1 activation.

## 1. INTRODUCTION

Placenta accreta spectrum (PAS) is defined as abnormal adherence of placental villi to the uterine myometrium and represents a major obstetric complication associated with life-threatening hemorrhage and maternal mortality. With increasing cesarean section rates, the incidence of PAS has increased from 0.8 per 1,000 deliveries in the 1980s to approximately 3 per 1,000 deliveries more recently [1, 2]. The spectrum can be divided into three categories: placenta accreta vera (accreta vera; attachment of placental villi to the myometrium without sufficient decidua), placenta increta (increta; deep invasion of placental villi into the myometrium), and placenta percreta (percreta; deep invasion of placental villi through the myometrium and uterine serosa) [1, 3]. Although antepartum diagnosis by ultrasonography and MRI has improved the accuracy of diagnosing increta and percreta, it remains difficult to predict all PAS cases that cause massive hemorrhage during delivery [3, 4]. The molecular biology involved in PAS pathogenesis is assumed to resemble tumor growth processes, such as angiogenesis and increased cellular invasiveness [3]. A single-cell transcriptome analysis in PAS revealed contributions of endothelial and decidual cells through alterations in the extracellular matrix and growth factors. Therefore, it remains controversial whether PAS is caused primarily by trophoblast invasion or by transcriptional changes in stromal cells such as endothelial and decidual cells [5].

Serine proteases such as thrombin and trypsin stimulate various cell types via G-protein-coupled protease-activated receptors (PARs) [6–8]. PARs comprise four subtypes: PAR-1, PAR-2, PAR-3, and PAR-4. PARs are expressed in a variety of normal human organs, and their activation regulates multiple physiological functions, including cell proliferation and migration, platelet aggregation, and inflammation [9]. Serine proteases cleave the extracellular N-terminal domain of PARs, generating a new N-terminus that acts as a tethered ligand and initiates signaling cascades leading to diverse cellular actions [9, 10]. Among PARs, PAR-1 and PAR-2 have been implicated in cancer cell proliferation and invasion, and many studies have described associations with clinical stage and poor prognosis [11–14]. PAR-1 has also been reported to be involved in early human placental development in the first trimester [15–17]. Furthermore, Erez et al. [18] reported a significant association between preeclampsia and increased PAR-1 expression in the human placenta. Several reports have suggested PAR-1 as a therapeutic target for preeclampsia [19–21]. However, PAR-1 expression and its role in PAS have not yet been established.

Thrombin, a PAR-1 agonist, contributes to platelet aggregation and fibrin formation. Platelets and fibrin are associated with maternal vascular remodeling and placental development [22–24]. Increased levels of plasma D-dimer and fibrin degradation products before delivery have been associated with FIGO grade in clinically and/or pathologically confirmed PAS [25]. A pathological study found excessive fibrinoid deposition at the uteroplacental interface in placenta accreta [26]. These findings suggest that fibrin formation and degradation occur in PAS placenta. However, molecular identification of fibrin formation and platelet aggregation in PAS has not been reported.

Therefore, in this study, we examined whether PAR-1 expression, platelet aggregation, and fibrin deposition in PAS differ from normal placenta and whether these findings differ according to the degree of PAS using paraffin-embedded specimens.

## 2. MATERIALS AND METHODS

### 2.1. Patients and tissue samples

Forty-nine cases that were histologically consistent with PAS were included in the study. For comparison of clinicopathologic factors and expression of PAR-1, platelet glycoprotein (GP) IIb/IIIa, and fibrin, 33 placentas with no apparent pathological changes were also included as controls. All cases were diagnosed at the Department of Pathology, University of Miyazaki, between 2006 and 2020. This study was approved by the institutional review boards of the University of Miyazaki (O-1785).

The definitive diagnosis of PAS was determined based on a combination of clinical imaging findings and gross and histological findings of tissue specimens submitted for pathological diagnosis. Placental and uterine tissue specimens were fixed in 10% buffered formalin after macroscopic examination of the cut surface. Then, for each case after fixation, 5–10 samples were taken from areas where gross examination suggested PAS, and paraffin blocks were prepared. In control cases, 4–5 specimens were taken randomly from the umbilical cord insertion site and the marginal region of the placenta. All sections (5 µm) were stained with hematoxylin and eosin (H&E). Two pathologists reviewed all sections by microscopy and made the histological diagnosis according to the diagnostic criteria of the College of American Pathologists conference [27]. The histologic diagnosis of PAS is defined as loss of decidua between placental villi and myometrium, resulting in direct adhesion of villi to the myometrium (Supplementary Figure 1). PAS was divided into three categories: accreta vera was defined as adhesion of placental villi to the superficial layer of the myometrium (31 cases); increta was defined as invasion of villi into the myometrium with thinning of the myometrium (8 cases); and percreta was defined as deep invasion into the myometrium with penetration of the serosa (10 cases). In the accreta vera group, additional confirmation was obtained in some cases by immunohistochemistry for desmin to demonstrate direct attachment of placental villi to myometrium. A retrospective review of clinicopathologic factors was performed. Maternal characteristics included age, gestational age at delivery, previous cesarean section, history of dilatation and curettage, whether the current pregnancy was achieved using assisted reproductive technology, and total blood loss at delivery.

### 2.2. Immunohistochemical study

Representative 10% formalin-fixed, paraffin-embedded specimens showing the most diagnostic histological findings in each case were selected for immunostaining. Sections (3 µm thick) were prepared, deparaffinized with xylene, and rehydrated through ethanol. Endogenous peroxidase activity was blocked with methanol containing 0.3% hydrogen peroxide. Sections were microwaved in 10 mM sodium citrate buffer (pH 6.0) containing Tween for 20 minutes for antigen retrieval.

All sections were immunohistochemically stained using antibodies against PAR-1 (Anti-F2R, mouse monoclonal; Santa Cruz Biotechnology, Santa Cruz, CA, USA), GPIIb/IIIa (sheep polyclonal; Affinity Biologicals, Inc., Hamilton, CA, USA), and fibrin (mouse monoclonal, clone 59D8; EMD Millipore Corp., Burlington, MA, USA). Additionally, myometrial tissue in accreta vera cases was evaluated by immunostaining with anti-desmin (rabbit polyclonal; Abcam, Cambridge, MA, USA). Sections were stained with EnVision anti-mouse or anti-rabbit (DAKO/ Agilent, Santa Clara, CA, USA) or an anti-sheep secondary antibody (Jackson ImmunoResearch, West Grove, PA, USA), according to the manufacturers’ instructions. Horseradish peroxidase activity was visualized using 3,3’-diaminobenzidine solution with hydrogen peroxide, and sections were counterstained with Mayer hematoxylin.

### 2.3. Scoring and quantification of immunohistochemistry

PAR-1 expression was evaluated in placental villi in both PAS and control groups. As an endogenous positive control, PAR-1 immunoreactivity was confirmed in the myometrium, decidualized stromal tissue, and vascular smooth muscle in the placenta and gestational uterus. Immunoreactivity scoring of PAR-1 expression was evaluated according to the method reported by Allred et al. [28], originally developed for assessment of estrogen and progesterone receptors in breast cancer.

The proportion of PAR-1-positive cells in placental villi was scored as follows: PS0, 0%; PS1, >0% to ≤1%; PS2, >1% to ≤10%; PS3, >10% to ≤33%; PS4, >33% to ≤67%; and PS5, >67% to ≤100%. Staining intensity was scored as follows: IS0, negative; IS1, weak; IS2, intermediate; and IS3, strong. If staining intensity was comparable to the endogenous positive control, the intensity score was assessed as IS2. The total score was calculated as the sum of the proportion score and intensity score.

Immunopositive areas for GPIIb/IIIa and fibrin were semi-quantified in the two fields of view with the largest positive areas under a 2× objective lens using WinRoof color image analysis software (Mitani, Fukui, Japan) [29]. Immunopositive areas were extracted as green areas using specific protocols based on color parameters (hue, lightness, and saturation). These areas were expressed as the immunopositive area ratio for each image.

### 2.4. Immunofluorescence

To identify PAR-1-expressing cells, we performed double immunofluorescence using antibodies against PAR-1 (anti-F2R, mouse monoclonal; Santa Cruz Biotechnology, Santa Cruz, CA, USA) and either HAI-1 (a cytotrophoblast marker; Abcam Japan, Tokyo, Japan) [30] or GPIIb/IIIa (sheep polyclonal; Affinity Biologicals, Ontario, Canada). CF488-conjugated donkey anti-mouse IgG (Biotium, Hayward, CA, USA) and CF568-conjugated donkey anti-rabbit IgG (Biotium, Hayward, CA, USA) or CF568-conjugated donkey anti-sheep IgG (Biotium, Hayward, CA, USA) were used as secondary antibodies. 4′,6-diamidino-2-phenylindole (DAPI) was used as a nuclear stain. Images were captured using spinning-disk confocal microscopy (SpinSR10; Evident, Tokyo, Japan) with a 40× objective lens. Merged fluorescence images were generated using image analysis software (cellSens; Evident, Tokyo, Japan).

### 2.5. Statistical analysis

Normality was assessed using the Shapiro–Wilk test. Data are presented as medians and ranges, box-and-whisker plots, or individual dot plots. Differences among groups were tested using the Kruskal–Wallis test with Dunn’s multiple-comparison test. Contingency analyses were performed using Pearson’s chi-square test. Relationships between variables were evaluated using Spearman’s rank correlation coefficient. Statistical analyses were performed using GraphPad Prism 8.4.3 (GraphPad Software Inc. San Diego, CA, USA). Statistical significance was set at *P* < 0.05.

## 3. RESULTS

### 3.1. Clinical characteristics of the study population

Table 1 shows clinical findings in the control and PAS groups. There were significant differences in maternal age at delivery and gestational week among groups. The frequency of previous cesarean section was higher in the increta and percreta groups than in the control and accreta vera groups. The frequency of assisted reproductive technology in the current pregnancy was higher in the accreta vera group than in the other groups. Total blood loss at delivery was significantly higher in all PAS groups than in the control group (*P* < 0.001) and was significantly higher in the percreta and increta groups than in the accreta vera group (*P* < 0.001).

**Table 1.**
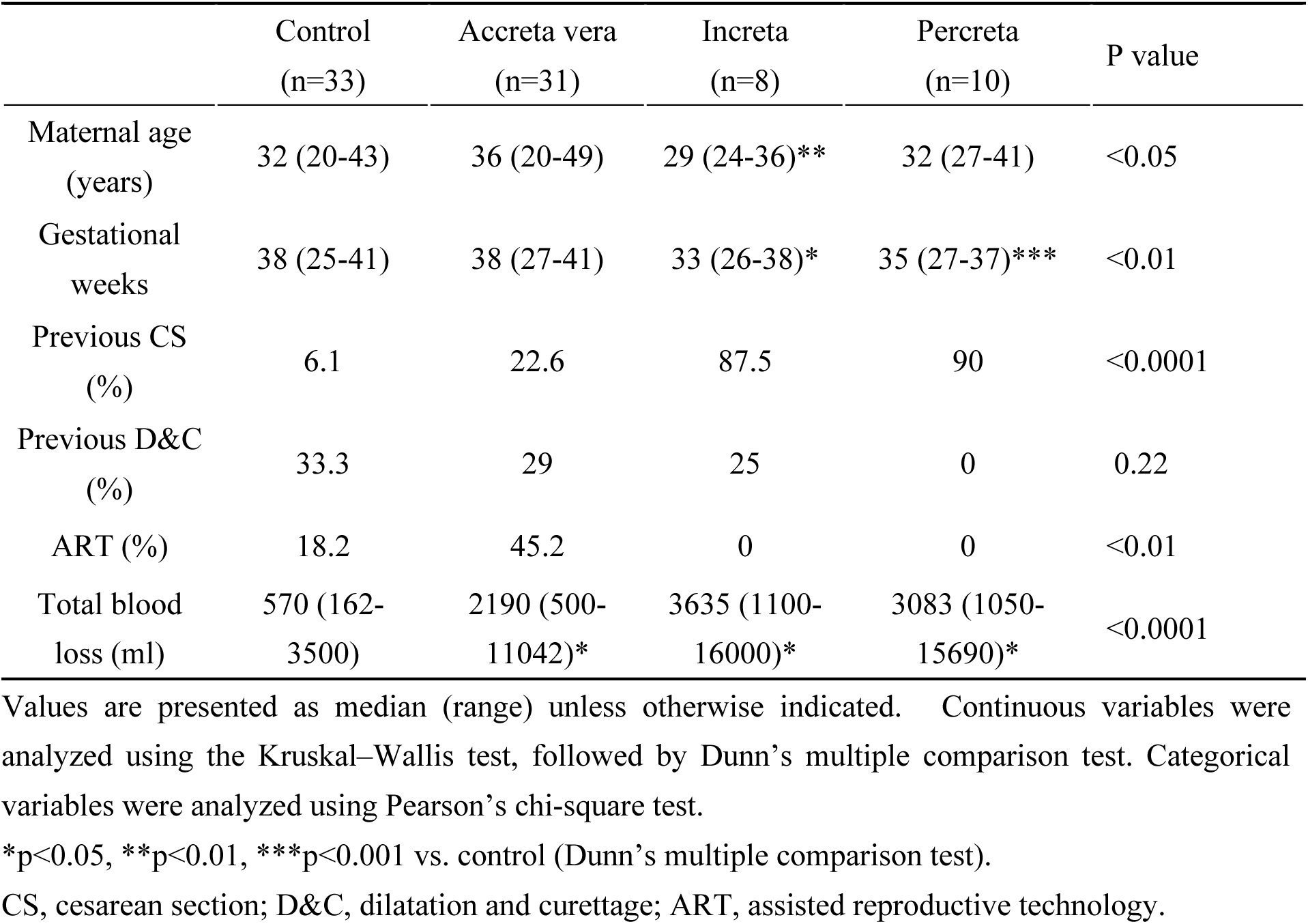
Clinical characteristics of control and placenta accreta spectrum groups.

|  | Control<br>(n=33) | Accreta vera<br>(n=31) | Increta<br>(n=8) | Percreta<br>(n=10) | P value |
| --- | --- | --- | --- | --- | --- |
| Maternal age<br>(years) | 32 (20-43) | 36 (20-49) | 29 (24-36)** | 32 (27-41) | <0.05 |
| Gestational<br>weeks | 38 (25-41) | 38 (27-41) | 33 (26-38)* | 35 (27-37)*** | <0.01 |
| Previous CS<br>(%) | 6.1 | 22.6 | 87.5 | 90 | <0.0001 |
| Previous D&C<br>(%) | 33.3 | 29 | 25 | 0 | 0.22 |
| ART (%) | 18.2 | 45.2 | 0 | 0 | <0.01 |
| Total blood<br>loss (ml) | 570 (162-3500) | 2190 (500-11042)* | 3635 (1100-16000)* | 3083 (1050-15690)* | <0.0001 |

Values are presented as median (range) unless otherwise indicated. Continuous variables were analyzed using the Kruskal–Wallis test, followed by Dunn’s multiple comparison test. Categorical variables were analyzed using Pearson’s chi-square test.
\*p<0.05, \*\*p<0.01, \*\*\*p<0.001 vs. control (Dunn’s multiple comparison test).
CS, cesarean section; D&C, dilatation and curettage; ART, assisted reproductive technology.

### 3.2. Representative microscopic findings of PAS

Representative microscopic findings on H&E staining in each PAS group and a representative desmin immunohistochemical finding in the accreta vera group are shown in Figure 1. In all cases diagnosed as PAS, placental villi were directly attached to the muscle layer without intervening decidual tissue. An intervening fibrinoid layer and some extravillous trophoblastic cells were occasionally observed between placental villi and myometrium. In the accreta vera group, placental villi were in direct contact with the myometrium, but there was no villous infiltration into the myometrium (Figure 1a). Immunohistochemistry for desmin, a smooth muscle marker, clearly highlighted the myometrium and revealed placental villi immediately adjacent to it (Figure 1b). Villous invasion into the myometrium and thinning of the myometrium were evident in the increta group (Figure 1c). In the percreta group, villous invasion into the myometrium was extensive and penetrated the uterine serosa (Figure 1d).

**Figure 1.**
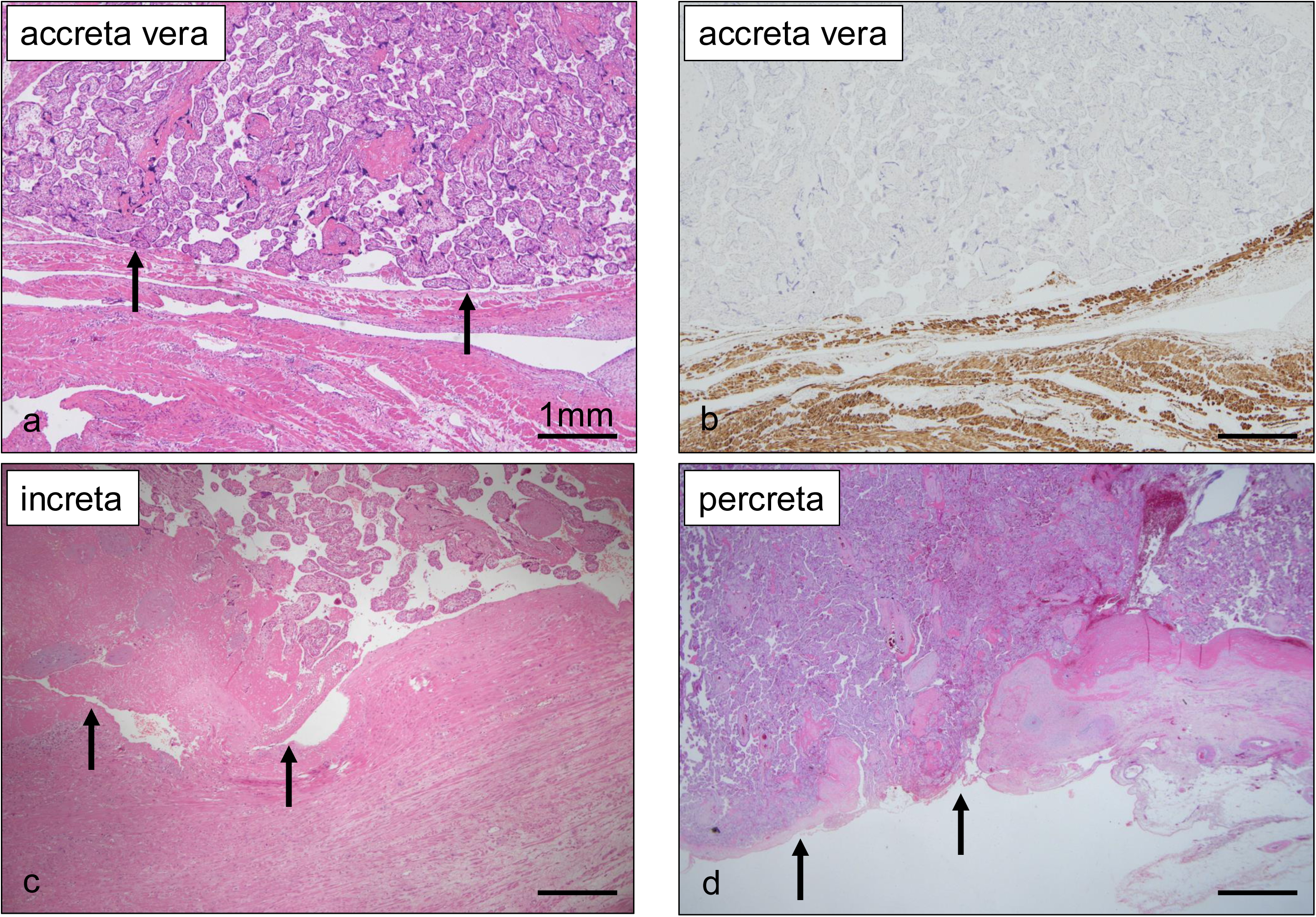
Representative microscopic and immunohistochemical findings of placenta accreta spectrum (accreta vera, increta, percreta). **a.** In accreta vera, placental villi are directly attached to the myometrium without intervening decidua (arrows). **b.** The myometrium is confirmed by immunohistochemistry for desmin. **c.** In increta, placental villi invade deep into the myometrium (arrows). **d.** In percreta, placental villi penetrate through the myometrium and uterine serosa (arrows); the serosal adipose tissue has disappeared.

### 3.3. PAR-1 expression in control and PAS groups

Representative features of PAR-1 expression in placental villi are shown in Figure 2a. In the control and accreta vera groups, the intensity and extent of PAR-1 expression varied among cases, and several intravillous cells expressed PAR-1. In the increta and percreta groups, PAR-1 expression was identified continuously or discontinuously in intravillous cells (Figure 2a). Immunofluorescence confirmed that HAI-1-positive cytotrophoblasts express PAR-1 in placental villi (Figure 2b) and that aggregated platelets in intervillous spaces also express PAR-1 (Figure 2c). In the increta and percreta groups, PAR-1 was moderately to strongly expressed in many cytotrophoblasts, particularly in the percreta group, where PAR-1 expression was observed in almost all cytotrophoblasts (Figure 2a).

**Figure 2.**
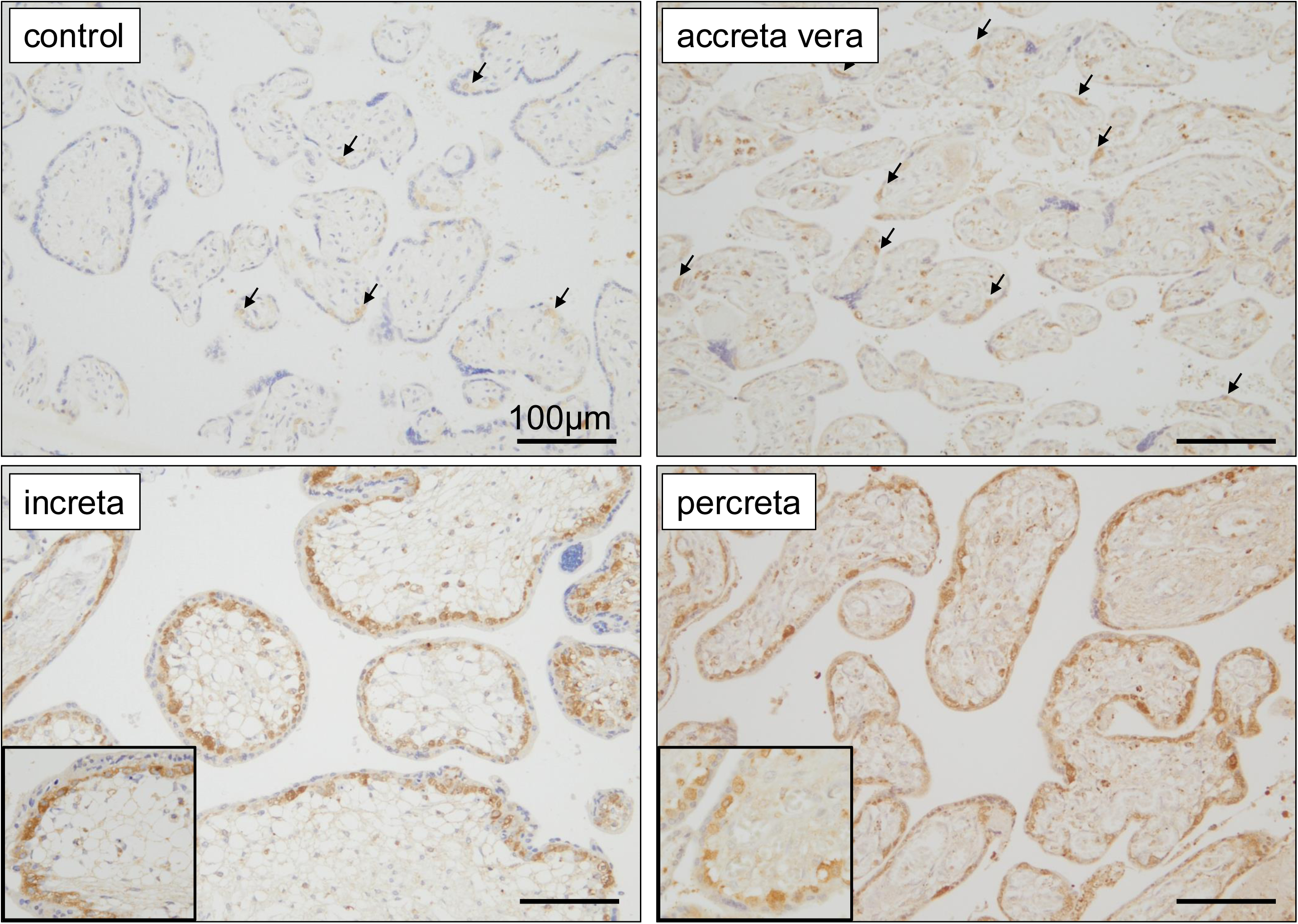
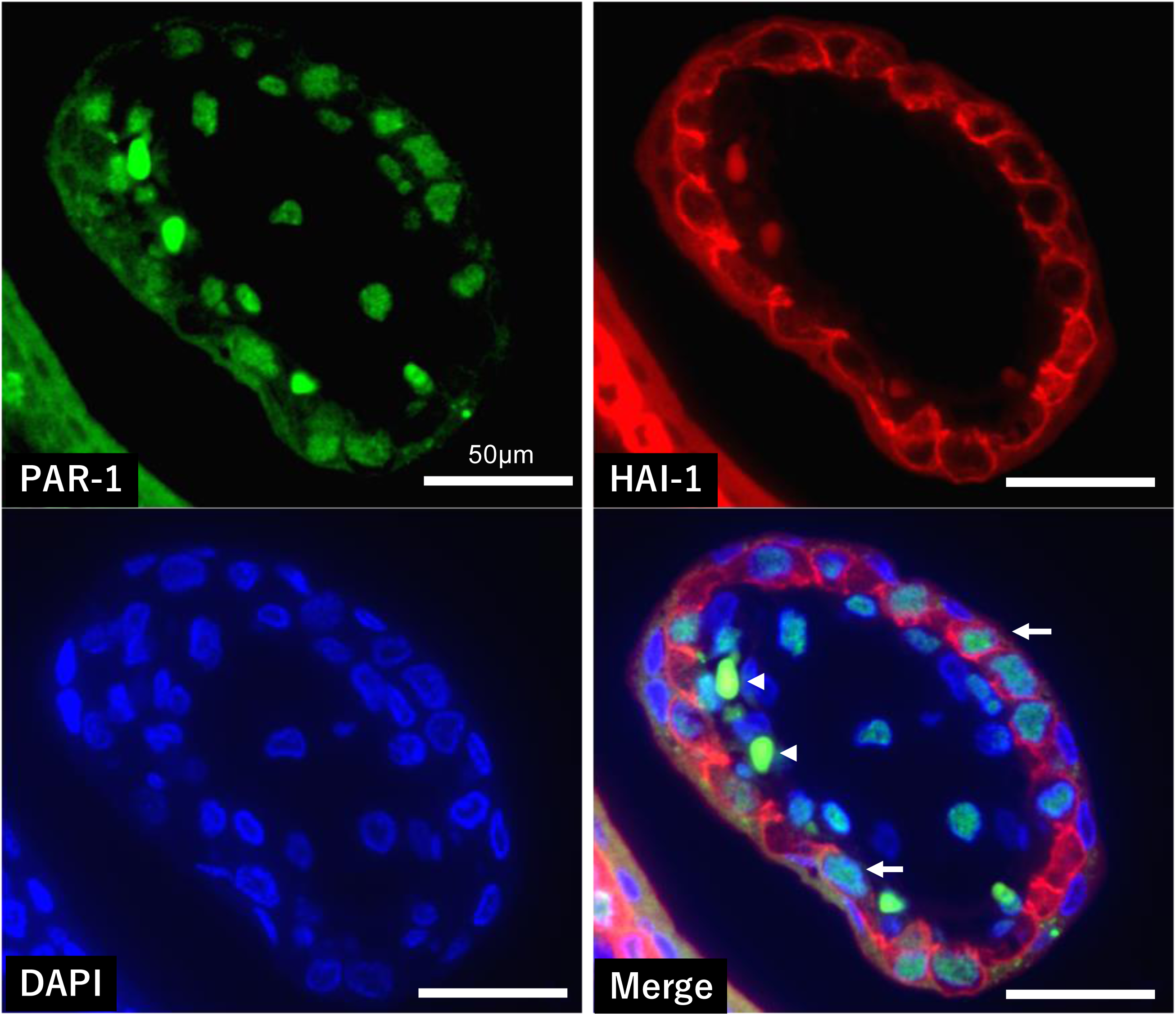
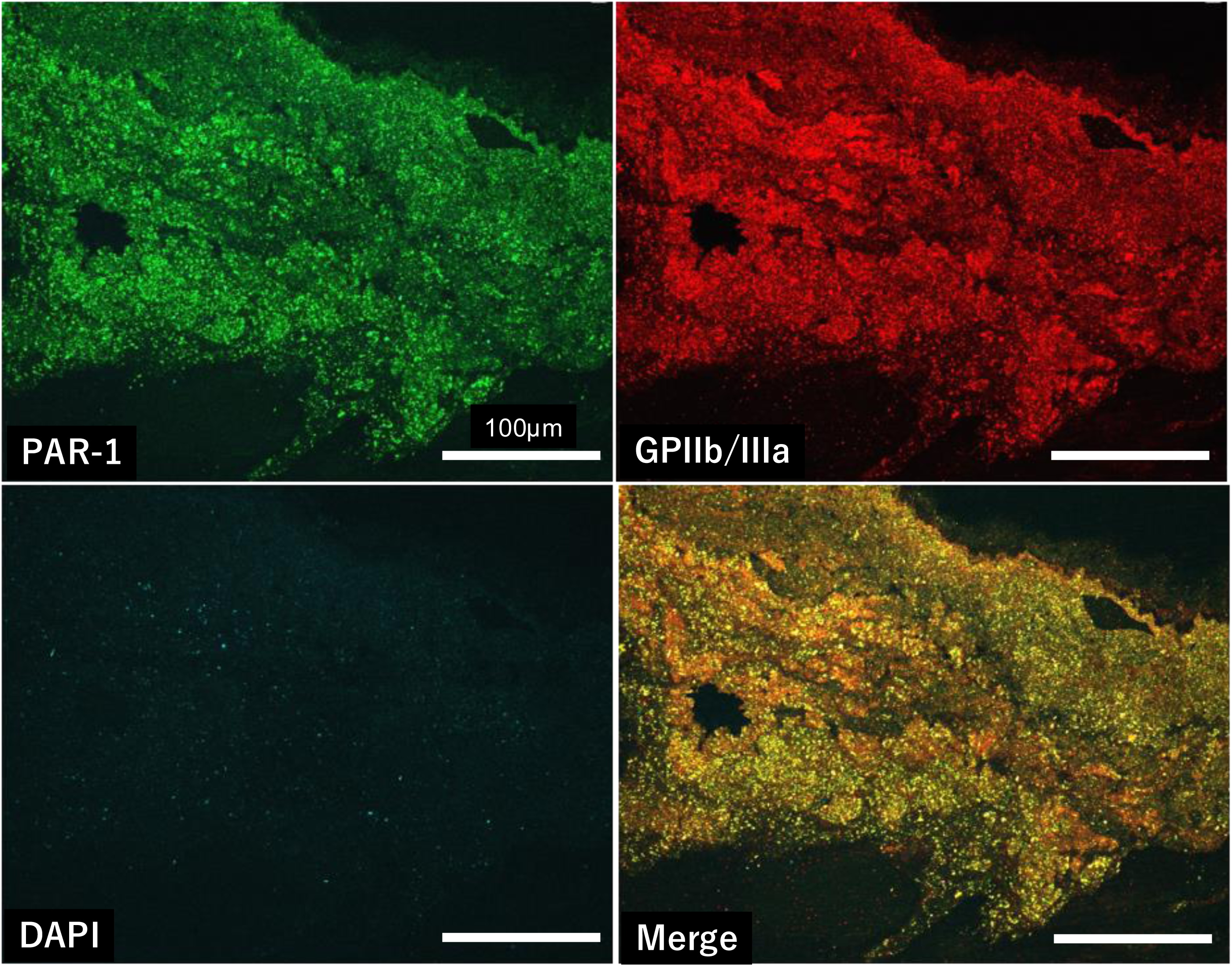
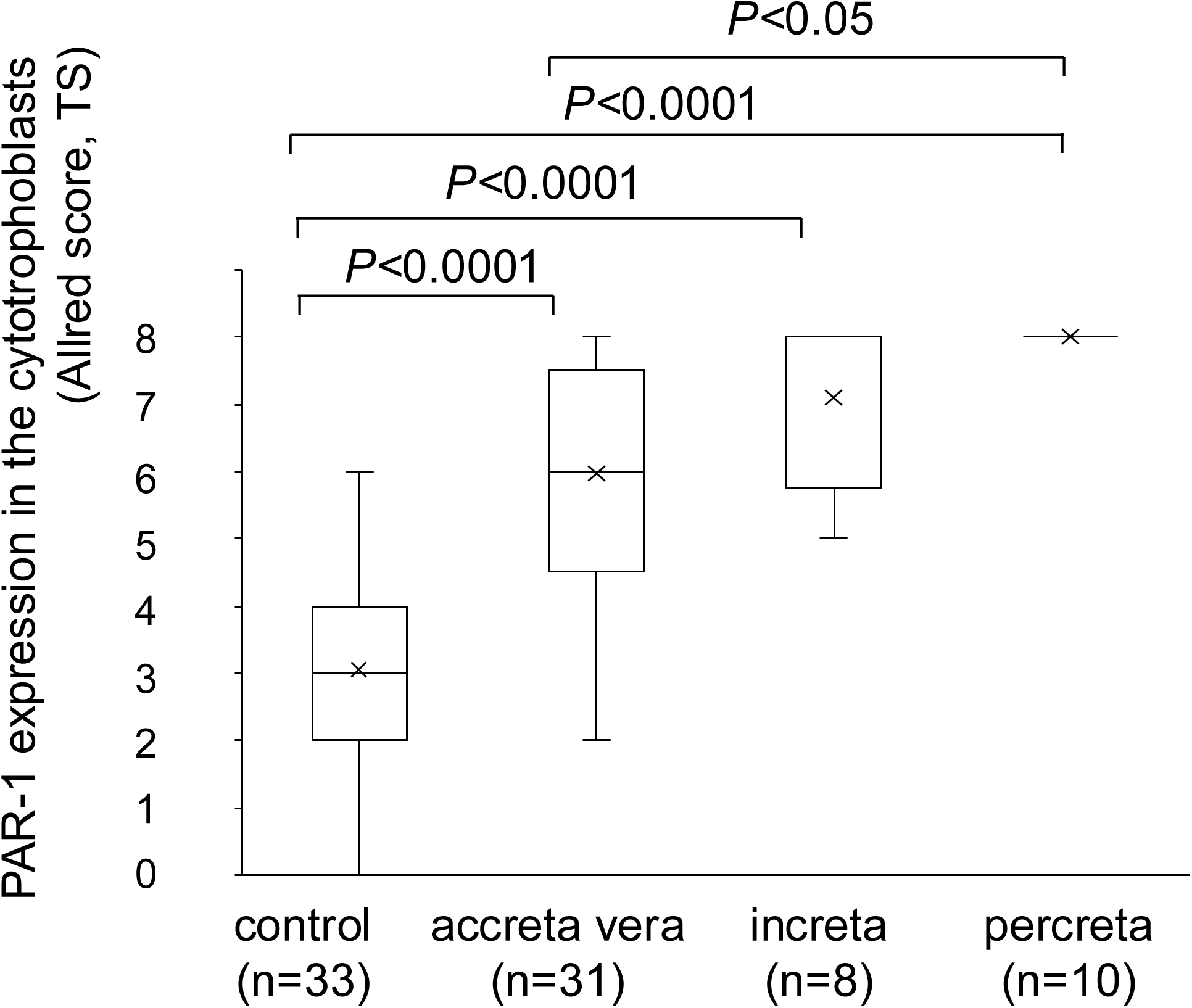
Representative immunohistochemical findings of PAR-1 expression in the control and PAS groups. **a.** In the control group, only a small number of intravillous cells are weakly positive for PAR-1 (arrows), scored as PS2 + IS1 = TS3. In accreta vera, PAR-1 immunopositivity is scored as PS4 + IS2 = TS6 (arrows). In increta and percreta, intravillous cells are strongly positive for PAR-1. In placental villi, the inner single-cell layer of cytotrophoblasts, but not the outer syncytiotrophoblasts, is immunopositive for PAR-1 (insets). PAR-1 immunopositivity is scored as PS5 + IS3 = TS8. **b.** Double immunofluorescence for PAR-1 and HAI-1 (a cytotrophoblast marker) with DAPI in placental villi (arrows). Arrowheads indicate erythrocyte autofluorescence. **c.** Double immunofluorescence for PAR-1 and GPIIb/IIIa (a platelet marker) with DAPI in the intervillous space. **d.** PAR-1 expression in cytotrophoblasts (Allred total score, TS) in the control and PAS groups. Statistical analysis was performed using the Kruskal–Wallis test with Dunn’s multiple-comparison test.

Immunoreactivity scoring of PAR-1 expression in the cytotrophoblasts was evaluated according to the method reported by Allred et al. [28]. Figure 2d shows Allred scores for PAR-1 expression in cytotrophoblasts in the control and PAS groups. In the percreta group, the Allred score for PAR-1 expression reached TS8, the maximum value. The score for PAR-1 expression increased with the degree of villous invasion (accreta vera, *P* < 0.0001; increta, *P* < 0.0001; percreta, *P* < 0.0001, vs control, respectively) and was significantly higher in the percreta group than in the accreta vera group (*P* < 0.05).

### 3.4. Immunohistochemical findings of platelet and fibrin in control placenta and PAS groups

Representative immunohistochemical findings for GPIIb/IIIa (a platelet marker) and fibrin are shown in Figure 3. In the control group, platelet- and fibrin-immunopositive areas were scattered in intervillous spaces. In the accreta vera group, GPIIb/IIIa- and fibrin-immunopositive areas were similarly scattered, and fibrin was also observed as thin bands between the myometrium and placental villi. In the increta and percreta groups, both platelet- and fibrin-immunopositive areas filled intervillous spaces, particularly in deeper areas (Figure 3).

**Figure 3.**
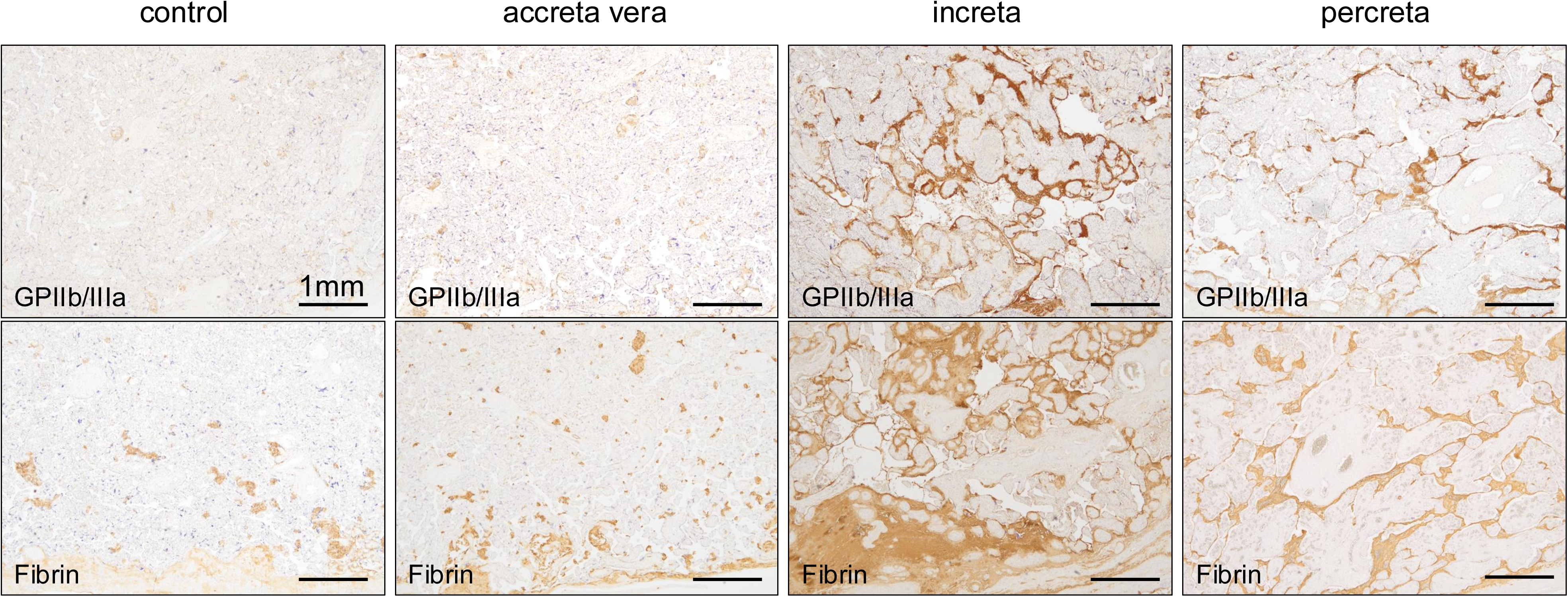
Representative immunohistochemical findings of GPIIb/IIIa and fibrin in placenta from the control and PAS groups. Platelets are highlighted by immunostaining for GPIIb/IIIa. Immunopositive areas for GPIIb/IIIa and fibrin are significantly larger in PAS groups than in the control group.

### 3.5. Increased platelet aggregation and fibrin deposition in the placenta in PAS groups

Figure 4 shows immunopositive areas for GPIIb/IIIa and fibrin in the control and PAS groups. The median value of immunopositive area for GPIIb/IIIa was larger in all PAS groups than in the control group (3.1%) (Figure 4a: accreta vera, 6.9%; increta, 10.9%; percreta, 7.9%; *P* < 0.0001 vs control for each). The median immunopositive area for fibrin was also larger in all PAS groups than in the control group (9.0%) (Figure 4b: accreta vera, 14.0%; increta, 28.5%; percreta, 29.1%; *P* < 0.0001 vs control, respectively).

**Figure 4.**
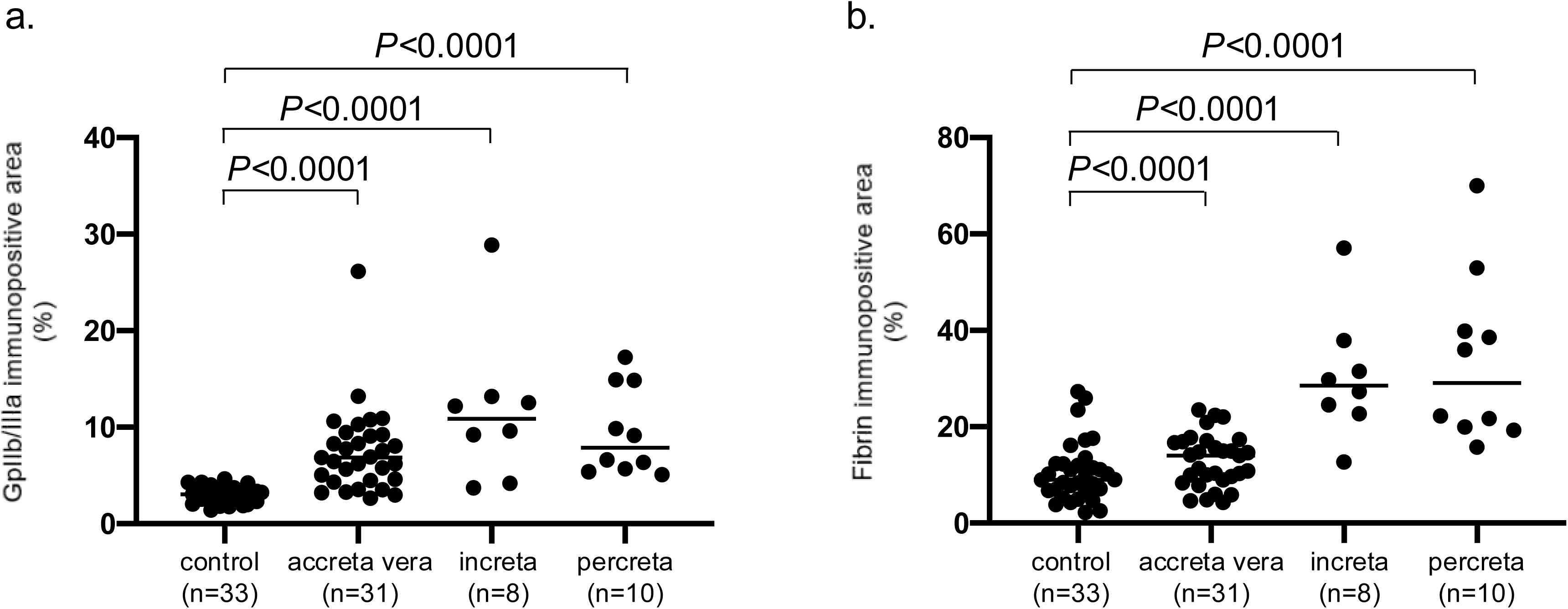
Comparison of immunopositive area ratios of GPIIb/IIIa and fibrin in the control and PAS groups (**a**: GPIIb/IIIa, **b**: fibrin). Statistical analysis was performed using the Kruskal–Wallis test with Dunn’s multiple-comparison test.

### 3.6. Relationships between degree of PAR-1 expression and platelet or fibrin deposition

Figure 5 shows relationships between Allred score for PAR-1 expression and immunopositive areas for GPIIb/IIIa and fibrin in the combined control and PAS cases. The immunopositive area for GPIIb/IIIa was positively correlated with the Allred score for PAR-1 expression (Figure 5a: r = 0.59, *P* < 0.0001). Fibrin immunopositive area also showed a positive, weak correlation with the Allred score for PAR-1 expression (Figure 5b: r = 0.34, *P* < 0.001).

**Figure 5.**
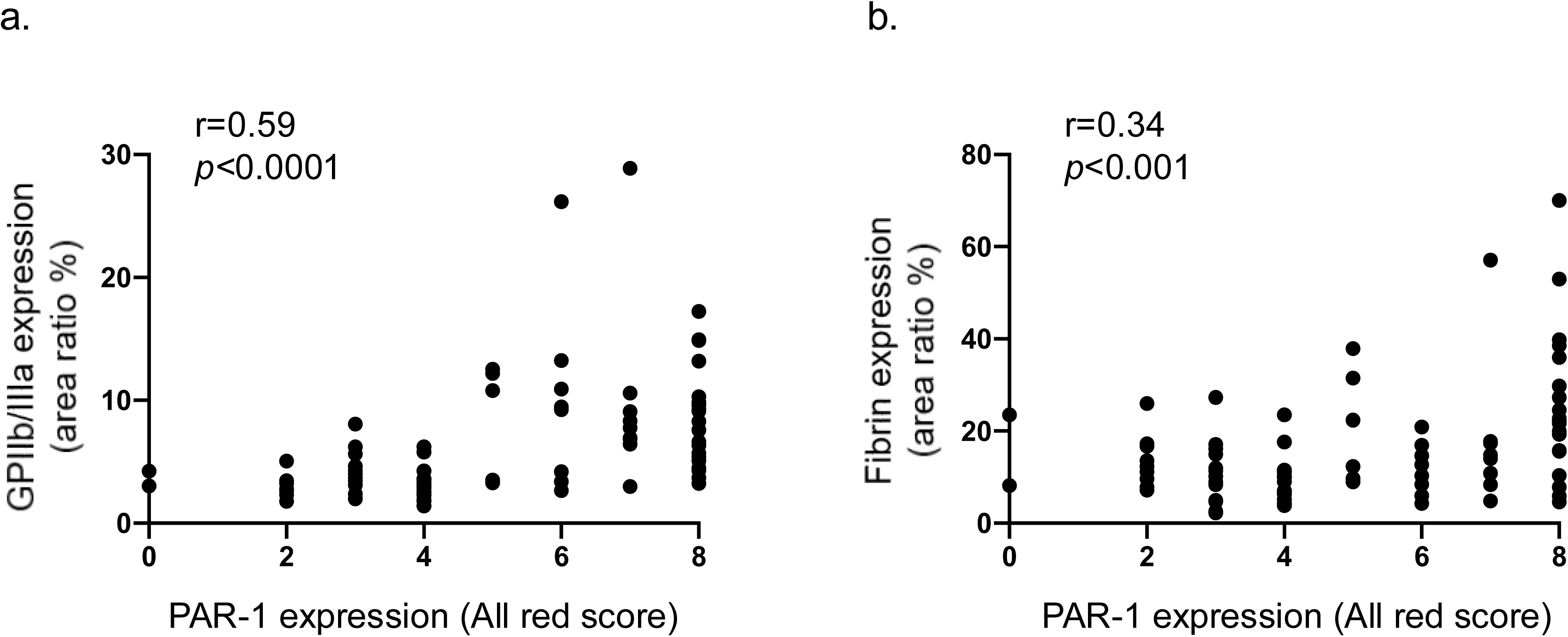
Correlation between PAR-1 expression and the area ratio of GPIIb/IIIa and fibrin. The horizontal axis shows the Allred score for PAR-1 expression across both control and PAS cases, whereas the vertical axis shows the immunopositive area ratio of GPIIb/IIIa (**a**) and fibrin (**b**). Statistical analysis was performed using Spearman’s rank correlation coefficient.

## 4. DISCUSSION

The present study showed that cytotrophoblasts and aggregated platelets express PAR-1 and that PAR-1 expression in placental villi increased with the degree of villous invasion in PAS. In addition, PAR-1 expression correlated with platelet aggregation and fibrin formation in the placenta. These observations are the first pathological evidence that increased PAR-1 expression and a thrombogenic placental condition are associated with the extent of villous invasion in PAS.

It is well known that a history of injury to the endometrium and myometrium, such as previous cesarean section, myomectomy, or endometrial curettage, is associated with PAS [31, 32]. Our previous study also showed that the accreta vera group had a high frequency of in vitro fertilization and that the increta or percreta group had a high frequency of previous cesarean section [33]. The previous and current studies suggest that risk factors differ between accreta vera and increta/percreta and that prior myometrial injury, rather than endometrial injury, is a risk factor for abnormal trophoblast invasion into the myometrium in increta and percreta.

In the present study, we focused on PAR-1, a member of the G-protein-coupled receptor family that plays key roles in hemostasis, thrombosis, inflammation, tumor growth processes, and placental implantation [34]. Activation of PAR-1 by proteases such as thrombin induces cellular effects including migration, proliferation, and extracellular matrix production [7]. During early development in human placental tissue, high mRNA and protein expression of PAR-1 and PAR-3 was found in cytotrophoblasts between 7 and 10 weeks’ gestation, whereas expression was absent at 12 weeks’ gestation and thereafter [15]. Low PAR-1 expression in third-trimester cytotrophoblasts from control placentas was comparable to that reported previously. PAR-1 expression is significantly higher in placenta with preeclampsia than in normal placenta [18]. Therefore, abnormal placental conditions in PAS may alter PAR-1 expression in the third trimester. Increased platelet aggregation and fibrin formation in PAS suggest enhanced thrombin generation in the diseased placenta. Thrombin induces IL-11 expression and secretion mainly via PAR-1 in human early gestational extravillous trophoblast cell lines and promotes extravillous trophoblast migration [16]. PAR-1-driven extravillous trophoblast migration is mediated via low-density lipoprotein receptor-related protein 5/6 [17]. High PAR-1 expression induces abnormal vascular remodeling and enhancement of extravillous trophoblast invasion [20, 21]. Taken together with our results, these reports suggest that PAR-1 may contribute to cytotrophoblast invasion in PAS and that thrombin is a candidate mediator of PAR-1 activation on cytotrophoblasts and of platelet aggregation.

PAR-1 can be upregulated by platelet-derived growth factor (PDGF), transforming growth factor (TGF)-β, thrombin, progesterone, and under conditions such as preeclampsia [7]. Platelets release PDGF and TGF-β upon activation [35, 36]. Aggregating human platelets stimulate PAR-1 expression in vascular smooth muscle cells through release of TGF-β1 and PDGFAB [37]. Activation of cell-surface PAR-1 by thrombin is itself a critical stimulus for PAR-1 gene expression in vascular endothelial cells via the Gi-linked Ras/MAPK signaling pathway [38]. In most increta and percreta cases, PAR-1 was diffusely and strongly expressed, suggesting not only local but also global environmental changes in these groups. Prior reports and the observed correlation between PAR-1 expression, platelet aggregation, and fibrin formation suggest that PDGF and thrombin itself may be involved in PAR-1 upregulation in PAS.

Proteins associated with the coagulation–fibrinolysis system, including antithrombin and components such as plasmin and plasminogen activator inhibitor-1, are dysregulated in sera from PAS patients compared with controls [39]. Increased levels of plasma D-dimer and fibrin degradation products before delivery have been associated with the degree of PAS [25]. These clinical studies suggest an imbalance in the coagulation–fibrinolytic system in PAS and support an enhanced thrombogenic condition, especially in increta and percreta groups, consistent with our findings. A pathological study of PAS showed dense, thick “fibrinoid” deposition between anchoring villi of the basal plate and the myometrium [26]. The present study confirmed this as fibrin deposition and found that deposition markedly increased in increta and percreta. In addition, fibrin deposition was accompanied by platelet aggregation, and platelets expressed PAR-1. These results suggest that fibrin is not simply deposited; rather, thrombin generated via the coagulation reaction may affect platelet and cytotrophoblast function via PAR-1 in increta and percreta.

This study has several limitations. First, the increta and percreta groups were small due to their rarity. Second, this histopathological study did not clarify the mechanisms underlying PAR-1 overexpression and the thrombogenic condition in PAS. Further basic studies are required to uncover the underlying mechanisms.

In conclusion, this study provides evidence that placentas from pregnancies complicated by PAS show increased PAR-1 expression in cytotrophoblasts and enhanced platelet aggregation and fibrin formation. These results suggest that PAR-1 expression is involved in PAS pathophysiology, including placental villous invasion, and that a thrombogenic placental condition may contribute to PAR-1 activation and overexpression.

## Data Availability

All data produced in the present study are available upon reasonable request to the authors.

## Acknowledgements

We thank Nahoko Udatsu and Kyoko Ohashi for their technical assistance. This study was supported by a Clinical Research Support Grant from the University of Miyazaki Hospital.

## Author contributions

Murasaki Aman designed and conducted the study, collected and analyzed the data, and wrote the manuscript. Toshihiro Gi and Nobuyuki Oguri also designed and conducted the study, collected and analyzed the data, and revised the manuscript. Eriko Nakamura, Kazunari Maekawa, Sayaka Moriguchi-Goto, Yuki Kodama, and Shinji Katsuragi participated in data analysis and manuscript revision. Yujiro Asada participated in the study design and revised the manuscript. Yuichiro Sato collected and analyzed the data and revised the manuscript. Atsushi Yamashita designed and supervised the study and revised the manuscript. All co-authors reviewed and approved the manuscript.

---

**Supplementary figure 1.**
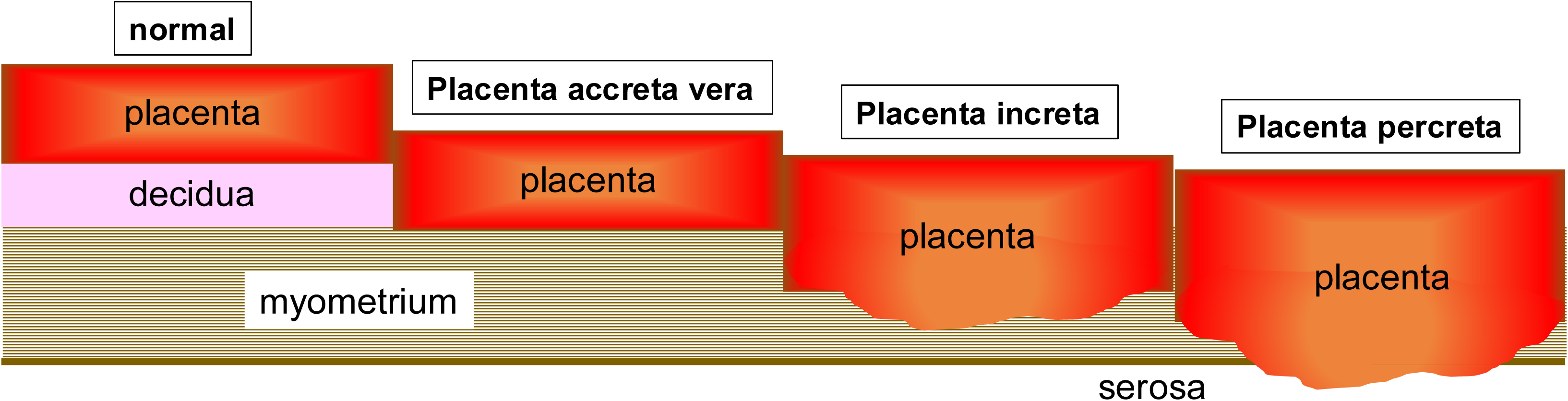
Definition of 3 categories of placenta accrete spectrum. Placenta accreta vera is defined as attachment of the placental villi to the myometrium without sufficient decidua. Placenta increta is defined as deep invasion of the placental villi into the myometrium. Placenta percreta is defined as deep invasion of the placental villi through the myometrium and uterine serosa.

